# Missing clinical trial data: the evidence gap in the safety of potential COVID-19 drugs

**DOI:** 10.1101/2020.05.30.20117523

**Authors:** Florence Rodgers, Toby Pepperrell, Sarai Keestra, Victoria Pilkington

**Author notes:** **Corresponding Author** Florence Rodgers.

## Abstract

**Background:** Several drugs are being repurposed for the treatment of the coronavirus disease 2019 (COVID-19) pandemic based on *in vitro* or early clinical findings. As these drugs are being used in varied regimens and dosages, it is important to enable synthesis of existing safety data from clinical trials. However, availability of safety information is limited by a lack of timely reporting of clinical trial results on public registries or through academic publication. We aimed to analyse the evidence gap in safety data by quantifying the number of missing clinical trial results for drugs potentially being repurposed for COVID-19 by conducting a rapid review of results posting on ClinicalTrials.gov and in academic publications.

**Methods:** ClinicalTrials.gov was searched for 19 drugs that have been identified as potential treatments for COVID-19. Relevant clinical trials for any prior indication were listed by identifier (NCT number) and checked for results and for timely result reporting (within 395 days of the primary completion date). Additionally, PubMed and Google Scholar were searched to identify publications of results not listed on the registry. A second, blinded search of 10% of trials was conducted to assess reviewer concordance.

**Results:** Of 3754 completed trials, 1516 (40.4%) did not post results on ClinicalTrials.gov or in the academic literature. 1172 (31.2%) completed trials had tabular results on ClinicalTrials.gov. A further 1066 (28.4%) completed trials had results from the literature search, but did not report results on ClinicalTrials.gov. Key drugs missing clinical trial results include hydroxychloroquine (37.0% completed trials unreported), favipiravir (77.8%) and lopinavir (40.5%).

**Conclusions:** There is an important evidence gap for the safety of drugs being repurposed for COVID-19. This uncertainty could cause a large burden of additional morbidity and mortality during the pandemic. We recommend caution in experimental drug use for non-severe disease and urge clinical trial sponsors to report missing results retrospectively.

## Background

Coronavirus disease 2019 (COVID-19) is a pandemic infection caused by severe acute respiratory syndrome coronavirus 2 (SARS-CoV-2). Its global spread has been rapid and unprecedented, at the time of writing 24.5 million confirmed cases have been reported with 832,378 deaths across 188 countries (1). Currently, treatment options for COVID-19 are limited. However, several drugs developed for other indications have shown promising results against SARS-CoV-2 in vitro, in animal models, or in compassionate use trials (2,3). Many of these drugs are now being experimentally repurposed for COVID-19 or are undergoing clinical trials in humans (4). Such candidates include nitazoxanide, remdesivir, favipiravir, lopinavir, darunavir, hydroxychloroquine, chloroquine, and ivermectin amongst others (5).

Investigations into the efficacy of these experimental treatments for COVID-19 are ongoing. Meanwhile, in response to positive media coverage, some speculative rather than evidence-based, governments are stockpiling vast supplies of these treatments in anticipation of their licensing for COVID-19. Furthermore, national regulatory institutions, meant to safeguard against the unsafe use of drugs, are under increasing pressure to relax approval standards to accelerate market-entry for COVID-19 treatments. For example, on April 27th 76 2020 the U.S. Food and Drug Administration (FDA) approved the antimalarial drug hydroxychloroquine for emergency treatment of COVID-19 with unknown optimal dosage and duration of treatment (6). However, the efficacy of hydroxychloroquine is still under question (7). Furthermore, it is cardiotoxic at the higher doses which may be indicated for COVID-19, causing QT prolongation leading to ventricular tachycardia and death (8). Care must be taken not to lose the rigorous safety standards usually stipulated for pharmaceuticals, even during a pandemic, to avoid unnecessary morbidity and mortality worldwide.

As many of these drugs have been used widely in other indications for years, there should be substantive information on safety, tolerability and pharmacokinetics available in the public domain, including public trial registries. Yet, whilst the International Committee of Medical Journal Editors (ICMJE) policy requires prospective registration of interventional studies on a WHO primary registry or on ClinicalTrials.gov, the ICMJE does not currently require researchers to report summary results on these registries before academic publication (9). However, according to the FDA Amendment Act 2007, the responsible party for some applicable clinical trials that are registered on ClinicalTrials.gov must report results to a public register within twelve months of the primary completion date, or in some cases risk a fine of $11,569 for every day results are delayed (10,11). Failure to share clinical trial results publicly can additionally have far-reaching consequences for health and public expenditure, as illustrated by the widespread stockpiling and prescription of oseltamivir (Tamiflu) during the swine flu outbreak in 2009, despite a lack of evidence on safety and efficacy (12). When the clinical study reports for this therapeutic were finally made publicly accessible, the benefits of the product turned out to be exaggerated and misrepresented in the journal publications compared to the underlying data. Following the same H1N1 swine flu outbreak a novel vaccine (Pandemrix) was rapidly rolled out, but it only emerged seven years later that the manufacturer had failed to disclose internal pharmacovigilance data showing narcolepsy to be a rare side-effect of recipients of the vaccine(13). Clinical trial transparency is therefore vital in order to maximise and unify the sharing of safety information and data on efficacy, during this pandemic and beyond.

Public clinical trial registries are an important tool for transparent collaborative research. On these registries, safety and efficacy data can be uploaded freely, shortly after completion of the study, and protocol and data collection methods are still quality assessed (14,15). In contrast, academic publication may stall for significant periods of time and can be costly and selective, with time-intensive writing and review processes. Furthermore, clinical trial registry data is available free of charge and can be pooled without concern for silent outcome switching or publication bias (16-18). Indeed, inclusion of unpublished study results from clinical trial registries in meta-analysis may provide important additional information on adverse events and more precise risk estimates than looking at journal publications alone (19). Moreover, academic publications often fail to disclose all information on adverse events occurring during clinical trials, and a systematic review comparing the journal publication with unpublished documents has suggested in 75% of the studies reviewed, the number of named adverse events was lower in published medical literature (20). This is especially important during the COVID-19 pandemic, as interest in potential treatments for COVID-19 generates even greater incentive than normal for the publication of studies with positive results only. Without rapid sharing of datasets for drugs that may be repurposed for COVID-19, secondary analyses of safety data will be arduous and often incomplete (14,21). This may slow down the biomedical innovation process and could lead to preventable side effects occurring in vulnerable patients if safety information remains missing. As the pharmaceutical pipeline is accelerated to address the COVID-19 pandemic, enhancing clinical trial transparency is more important than ever.

In this rapid review of ClinicalTrials.gov, we aimed to determine the scale of unpublished clinical trial results which may hinder safety reviews of repurposed drugs for COVID-19. We reviewed the number of completed or terminated trials that have not reported results for an extensive, list of medications being repurposed for COVID-19, looking at all previous indications for these drugs. Specifically, we searched for any trial results that have not been made available to the public, with no results published on either the ClinicalTrial.gov registry or in the academic literature.

## Methods

We selected 19 potential treatments for coronavirus diseases based on information found in potential COVID-19 treatment reviews (5,22,23). The drugs assessed were pirfenidone, hydroxychloroquine, azithromycin, favipiravir, oseltamivir, sarilumab, tocilizumab, remdesivir, leflunomide, interferon-alpha, lopinavir-ritonavir, darunavir-ritonavir, baloxivir marboxil, umifenovir, interferon-beta, sofosbuvir, nitazoxanide, APN01 and ivermectin (Table 1). Synonyms and chemical names for these drugs were taken from pubchem.ncbi.nlm.nih.gov (Appendix 1) (24).

**Table 1:** Trials registered to NCT for repurposed drugs, of which ongoing and suspended or withdrawn.

| Generic name | Total registered | Ongoing | Suspended/withdrawn |
| --- | --- | --- | --- |
| Pirfenidone | 86 | 30 (34.9%) | 2 (2.3%) |
| Hydroxychloroquine sulfate | 233 | 74 (31.8%) | 5 (2.1%) |
| Azithromycin | 457 | 98 (21.4%) | 16 (3.5%) |
| Favipiravir | 12 | 3 (25.0%) | 0 (0.0%) |
| Oseltamivir | 127 | 18 (14.2%) | 8 (6.3%) |
| Sarilumab | 37 | 19 (51.4%) | 0 (0.0%) |
| Tocilizumab (atlizumab) | 360 | 84 (23.3%) | 9 (2.5%) |
| Remdesivir | 12 | 11 (91.7%) | 0 (0.0%) |
| Leflunomide | 25 | 4 (16.0%) | 1 (4.0%) |
| Interferon-alpha | 1161 | 71 (6.1%) | 41 (3.5%) |
| Lopinavir / ritonavir | 1015 | 66 (6.5%) | 22 (2.2%) |
| Darunavir / ritonavir | 246 | 26 (10.6%) | 4 (1.6%) |
| Baloxivir marboxil | 9 | 4 (44.4%) | 0 (0.0%) |
| Umifenovir | 10 | 8 (80.0%) | 0 (0.0%) |
| Interferon-beta | 343 | 43 (12.5%) | 15 (4.4%) |
| Sofosbuvir | 261 | 75 (28.7%) | 10 (3.8%) |
| Nitazoxanide | 57 | 8 (14.0%) | 1 (1.8%) |
| APN01 (ACE2 analogue) | 1 | 0 (0.0%) | 0 (0.0%) |
| Ivermectin | 101 | 19 (18.8%) | 4 (4.0%) |
| **TOTALS** | **4553** | **661 (14.5%)** | **138 (3.0%)** |

The U.S. clinical trials registry (ClinicalTrials.gov), was searched for all trials that listed these drugs as an intervention. Results of the search were downloaded on 4^th^ April 2020 (25). Numbers of trials with and without results on the registry were recorded. For trials without results, trial status was determined (completed, ongoing, suspended, terminated or withdrawn). Trials listing ‘primary completion date’ in the future were counted as ongoing, if no primary completion date was available then the study completion date was used. Listed trial status was used to identify terminated, suspended and withdrawn trials.

For all trials without results on ClinicalTrials.gov, a three-step process was followed between 4^th^ – 27^th^ April 2020 to determine whether results were reported elsewhere through academic publication (Figure 1).

1. Publications automatically indexed by clinical trial identifier (NCT number) on ClinicalTrials.gov were screened and included based on criteria below. If multiple publications were listed, the earliest dated publication was selected.
2. If results were not available on the registry, the NCT number was used to search and screen academic publications in PubMed.
3. If the PubMed search did not retrieve an academic publication, an additional search was conducted in Google Scholar using the following search terms in succession: clinical trial identifier; listed title; intervention name with primary investigator’s name. For each search, the first twenty results were screened.

**Figure 1:**
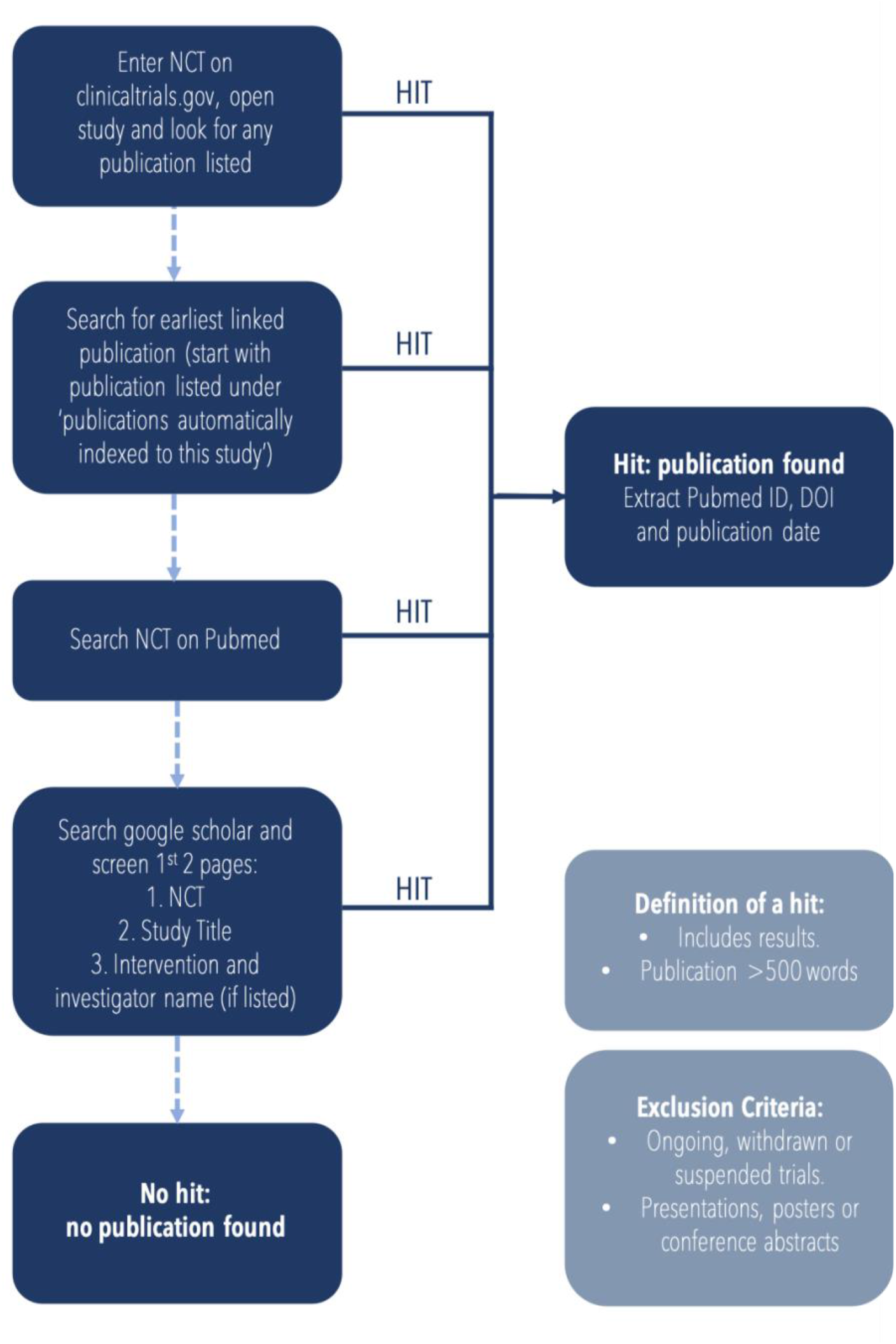
Flow chart depicting the methodology used to search for and identify relevant publications for each of the trials listed on CT.gov

If a publication did not include the clinical trial identifier, it was cross-referenced with the primary investigator, study design, intervention and outcomes listed on ClinicalTrials.gov to assess relevance. We excluded publications that had fewer than 500 words, as well as conference abstracts, posters, presentations, and non-English texts.

Publications of results were recorded by PMID, DOI, and publication date. Trials with results on ClinicalTrials.gov combined with those with a journal publication gave a total number of trials with results where results were found in the public domain. This allowed approximation of registered trials without results.

Additionally, overdue trials were calculated as any completed trial with no result on the registry and a primary completion date before 18^th^ April 2019, 395 days prior to final analysis (1 year + 30-day grace period). This is the standard outlined in the FDAAA 2007 and used as a reference throughout this study despite not all included trials being covered by the law (10). This is also consistent with international ethical standards for timely results dissemination set by the World Health Organisation (26).

A second review was conducted by a different researcher on 10% of trials for each drug to check concordance between reviewers. The protocol during the second review remained unchanged and researchers were blinded to the results of the first review. A random number generator was used to select trials for second review. Concordance was calculated by simple percent agreement on a results judgement; above 80% was deemed acceptable from consulting experts and in line with recommendations in the literature (27). Results published between the dates of first and second review (29^th^ April – 9^th^ May 2020) were not counted in the concordance.

## Results

Nineteen drugs were screened, encompassing a total of 4553 clinical trials registered on ClinicalTrials.gov (Table 1). We excluded 799, with 661 ongoing (primary completion date in the future) and 138 with a trial status of suspended or withdrawn. Figure 2 shows the number of trials found on ClinicalTrials.gov, those excluded from this analysis, and the final results status of all included trials. All recorded percentages in text are in relation to the 3754 completed trials, seen in Table 2.

**Figure 2:**
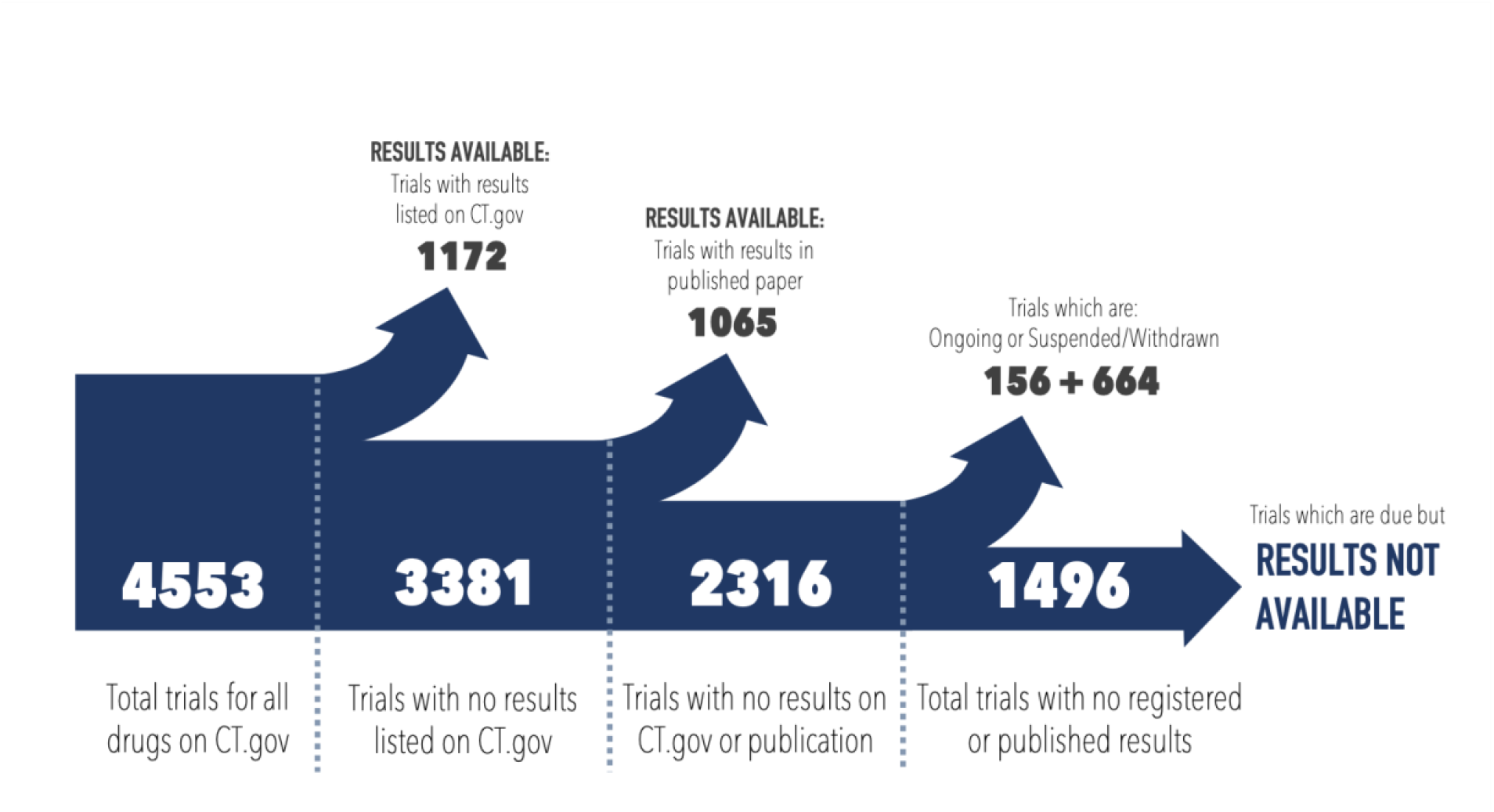
Flow diagram displaying the numbers of registered trials identified on CT.gov and the proportion of these which ultimately have no results available. The number of trials which had results available from various sources or had been withdrawn/suspended or trails for which results were not yet due are also shown.

**Table 2:**
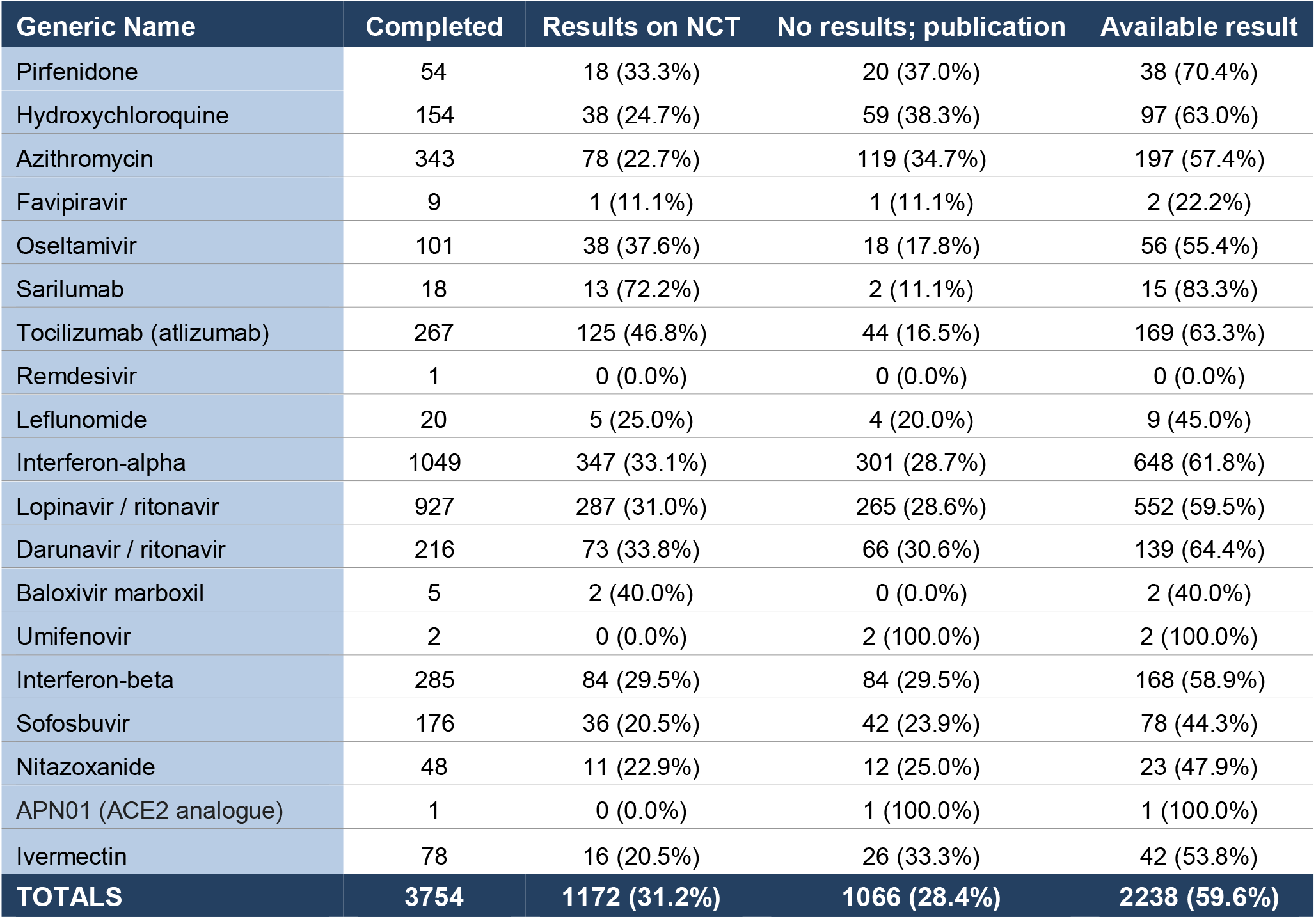
Number of completed trials registered on ClinicalTrials.gov, of which results have been published on ClinicalTrials.gov or in the academic literature.

| Generic Name | Completed | Results on NCT | No results; publication | Available result |
| --- | --- | --- | --- | --- |
| Pirfenidone | 54 | 18 (33.3%) | 20 (37.0%) | 38 (70.4%) |
| Hydroxychloroquine | 154 | 38 (24.7%) | 59 (38.3%) | 97 (63.0%) |
| Azithromycin | 343 | 78 (22.7%) | 119 (34.7%) | 197 (57.4%) |
| Favipiravir | 9 | 1 (11.1%) | 1 (11.1%) | 2 (22.2%) |
| Oseltamivir | 101 | 38 (37.6%) | 18 (17.8%) | 56 (55.4%) |
| Sarilumab | 18 | 13 (72.2%) | 2 (11.1%) | 15 (83.3%) |
| Tocilizumab (atlizumab) | 267 | 125 (46.8%) | 44 (16.5%) | 169 (63.3%) |
| Remdesivir | 1 | 0 (0.0%) | 0 (0.0%) | 0 (0.0%) |
| Leflunomide | 20 | 5 (25.0%) | 4 (20.0%) | 9 (45.0%) |
| Interferon-alpha | 1049 | 347 (33.1%) | 301 (28.7%) | 648 (61.8%) |
| Lopinavir / ritonavir | 927 | 287 (31.0%) | 265 (28.6%) | 552 (59.5%) |
| Darunavir / ritonavir | 216 | 73 (33.8%) | 66 (30.6%) | 139 (64.4%) |
| Baloxivir marboxil | 5 | 2 (40.0%) | 0 (0.0%) | 2 (40.0%) |
| Umifenovir | 2 | 0 (0.0%) | 2 (100.0%) | 2 (100.0%) |
| Interferon-beta | 285 | 84 (29.5%) | 84 (29.5%) | 168 (58.9%) |
| Sofosbuvir | 176 | 36 (20.5%) | 42 (23.9%) | 78 (44.3%) |
| Nitazoxanide | 48 | 11 (22.9%) | 12 (25.0%) | 23 (47.9%) |
| APN01 (ACE2 analogue) | 1 | 0 (0.0%) | 1 (100.0%) | 1 (100.0%) |
| Ivermectin | 78 | 16 (20.5%) | 26 (33.3%) | 42 (53.8%) |
| **TOTALS** | **3754** | **1172 (31.2%)** | **1066 (28.4%)** | **2238 (59.6%)** |

In sum, our protocol revealed 2238 (59.6%) completed trials had published results either on the registry or in the academic literature (Table 2). Of these, 1172 (31.2%) completed trials had tabular results on ClinicalTrials.gov (Table 2). A further 1066 (28.4%) completed trials had results from the literature search, but did not report results on ClinicalTrials.gov (Table 2). Across the 19 drugs which may be repurposed for the treatment of COVID-19, 1516 (40.4%) of completed clinical trials listed on ClinicalTrials.gov were missing results. Figure 3 shows the proportions of trial results available on ClinicalTrials.gov, available in the literature, and those with no results available.

**Figure 3:**
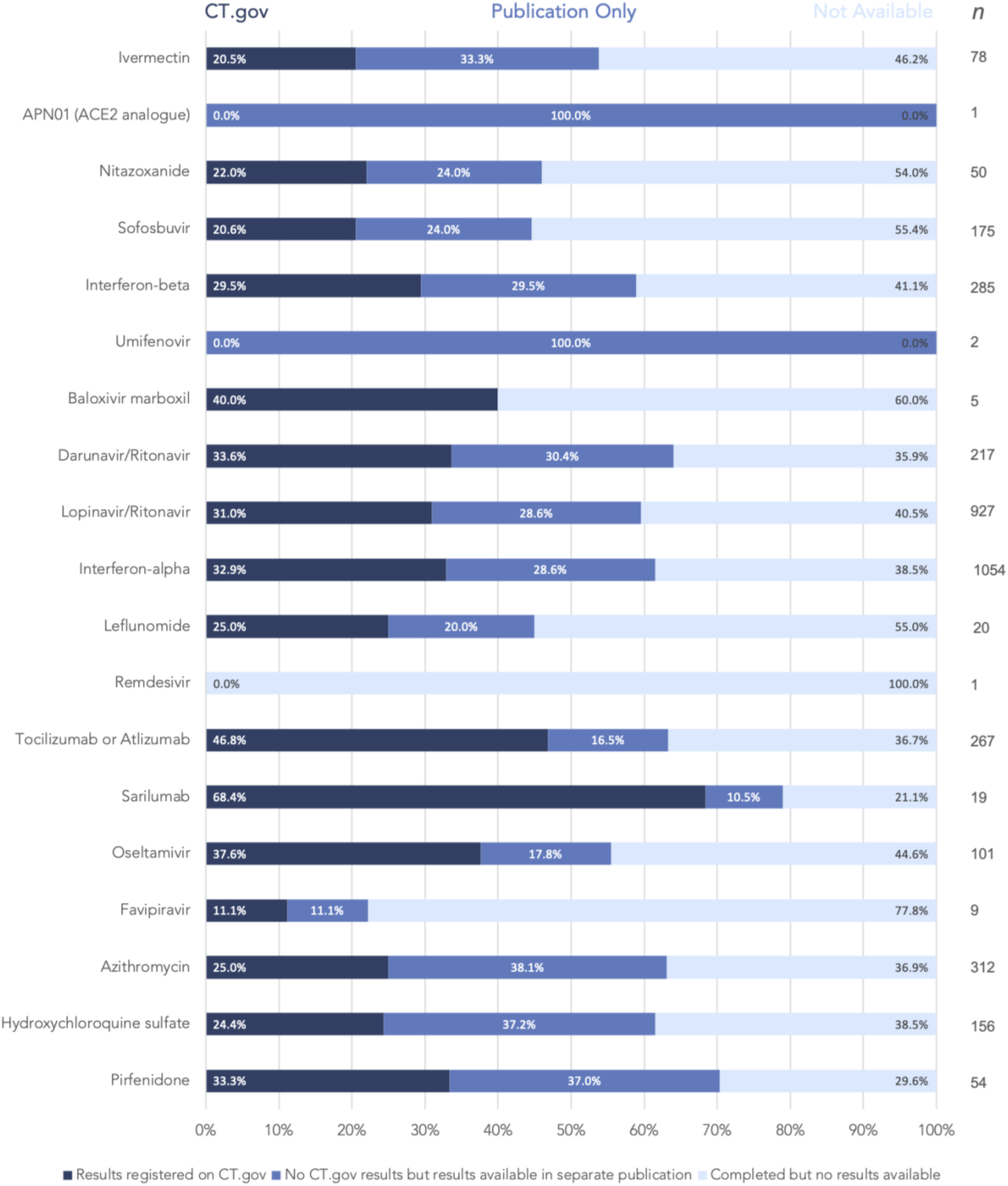
Bar chart displaying the proportion of trial results available across all potential COVID-19 interventions - categorised into those registered fully on CT.gov, those with results available elsewhere and those with no results available.

Of the 3754 completed studies, 2379 (63.4%) were without results on ClinicalTrials.gov outside of the 395-day timeframe mandated by the FDAAA 2007 (although not all trials may have been covered by the FDAAA 2007). 1008 (26.9%) of these had published results in the academic literature (10).

In the blinded second review of 341 (10%) trials, the same results status was found 83.6% of the time meeting our 80-100% threshold for acceptable concordance in searches.

## Discussion

Of the completed clinical trials for existing drugs that may be repurposed for COVID-19, 40.4% did not report results on either ClinicalTrials.gov or through academic publication (Table 2). This shows a large gap in the evidence base potentially regarding the adverse effects of these drugs, limiting attempts to comprehensively review their safety before potential global distribution for the COVID-19 pandemic. The 2238 (59.6%) completed studies with available results were comprised of 1172 (31.2%) with results on the registry and 1066 (28.4%) without results on the registry, but with results from a standardised search of the literature (Table 2). Furthermore, 2379 (63.4%) studies without registry results were outside of the 395 day timeframe for results publication as mandated by the FDAAA 2007 (10), although 1008 (26.9%) of these had results published in academic literature, they still failed to upload their results on ClinicalTrials.gov. Not all trials include in this study are covered by the FDAAA 2007, but the twelve month deadline remains an important benchmark for good scientific practice (26). With 40.4% of clinical trial results unavailable for potential COVID-19 treatments, the data for clinical decision making regarding the safety of these therapeutics are limited. If any drug with an incomplete evidence base is used during the pandemic, even in compassionate use programmes, there is a risk of avoidable harm being done because of missing adverse safety data. An evidence gap was revealed for drugs which have had extensive media coverage such as hydroxychloroquine (37.0% without results), favipiravir (77.8%) and lopinavir (40.5%) (28-30). These drugs are currently being used in COVID-19 patients and clinical trials across the globe, sometimes in novel regimens and doses (7,31,32). Clinicians currently have few treatment options available, but with greater transparency and proactiveness from trial sponsors regarding the posting of results, there would be less risk of unforeseen adverse outcomes, especially in the treatment of mild-moderate COVID-19 as in the PIONEER trial (31).

It is recognised that there are sources of safety data outside of clinical trial registries, for example pharmacovigilance registers, however these databases are not always free to access and publicly available. In contrast with clinical trial registries, there is often no obligation for all adverse events to be reported by trial sponsors. Public health decision-makers, guideline developers, clinicians, and patients therefore rely on clinical trial registries, systematic reviews, and meta-analyses to inform treatment decisions. Evidence gaps and publication bias therefore have the potential to influence clinical practice and drug usage worldwide, particularly in a treatment landscape as changeable as the COVID-19 pandemic. If clinical decisions are based on incomplete evidence, this can result in avoidable morbidity and mortality, especially if unsafe drugs or ineffective treatments are given on a large scale. Sponsors and researchers alike carry an ethical responsibility towards clinical trial participants, who consent to participate in research in order to contribute to scientific understanding and improved clinical practice, to make results publicly available (33).

Our study reveals an important evidence gap regarding existing pharmaceuticals potentially being repurposed for COVID-19. However, the proportion of studies with results available in the academic literature is an approximation and there are several limitations to our study. Firstly, our trial population was limited only to those registered on ClinicalTrials.gov. While ClinicalTrials.gov is the largest registry in the world, with over 340,000 registrations as of writing and an order of magnitude greater than the next largest journal, additional trials on these therapies may have been registered elsewhere. However, it is unlikely that these would be expected to report at a different rate than those registered to ClinicalTrials.gov. Secondly, our strategy for locating publications included only those listed on ClinicalTrials.gov and identified through searches on PubMed and Google Scholar, which are open-access resources that should cover a majority of published clinical research. While including proprietary databases like Scopus or Ovid may have located some additional publications, we do not believe this would have substantially impacted our overall results (34). Thirdly, we are aware that trials that were not registered at all or published in non-English language journals without inclusion of the NCT number would not have been captured by our methodology. Finally, searcher heterogeneity and difficulty identifying results publication in the academic literature limits accuracy in any manual publication search, however, our search strategy was standardised and produced a high level of agreement between assessors (83.6%) in a check of a 10% random sample. However, any discordance between reviewers only reveals the inherent difficulties in finding results for the drugs in question, especially if the trial identification number was not included in line with CONSORT standards (35).

Our findings add to the existing evidence of the dearth of clinical trial reporting on public registries. This analysis investigated clinical trials of existing drugs currently being considered for use for COVID-19. However, given the diversity of drug classes included in this report, findings are likely to be representative of many pharmaceuticals. This presents a major problem for researchers attempting to summarise safety and efficacy by pooling trial data (36). Since academic publications often summarise key findings only, secondary research efforts are impinged by the incomplete publishing of all trial outcomes. ClinicalTrials.gov, in contrast, provides a forum to share complete safety and efficacy data reports, as well as facilitating consistent data reporting in a timely manner (14,15). Prior research has shown that results reported to ClincialTrials.gov were often more complete, especially for safety data, when compared to matched journal publications (16-18). However, the availability of this data depends on researchers registering trials and uploading results in a timely manner, within twelve months of the primary completion date.

The International Committee of Medical Journal Editors (ICMJE) and the editorial offices of medical journals could play an important role in improving the lack of timely results posting on clinical trial registries by demanding submission of a link to summary results on public registries before academic publication, although this may mean that the publication bias of positive, ‘publishable’ results may trickle down to reporting on public registries as well. Furthermore, public funders and institutional publication funds could demand that trial sponsors post their results before allocating funding for academic (open-access) publication. These funding bodies could also deny individual sponsors funding if they have in the past violated clinical trial reporting rules (37). At the very least, journals should conform to the CONSORT statement in ensuring that registry identification numbers are clearly indicated in the abstract, full-text, and meta-data of published clinical trials in order promote discoverability and record linkage between registries and publications (35). Finally, clinical trial sponsors, such as universities, hospitals, public research institutions, and pharmaceutical companies, should themselves work towards improving their institutional clinical trial reporting performance by making use of available resources that provide detailed step-by-step instructions as to how to go about this task (11). Especially during the COVID-19 pandemic, it is of great importance that trials sponsors release summary results on these registries retrospectively to inform decision making around existing treatments being re-purposed for COVID-19.

## Conclusions

Overall, our findings reveal a significant evidence gap for the safety of drugs being repurposed for COVID-19. This uncertainty could cause a large burden of extra morbidity in the global pandemic. We recommend caution in experimental drug use for non-severe disease and urge trial sponsors to report missing results retrospectively. Medicine during the COVID-19 pandemic cannot be evidence-based if a large proportion of the evidence is missing.

**List of Abbreviations**
COVID-19: Corona Virus Disease 2019
CONSORT: Consolidated Standards of Reporting Trials
FDAAA2007: FDA Amendment Act 2007
ICMJE: International Committee of Medical Journal Editors
NCT Number: ClinicalTrials.gov identifier
PMID: PubMed ID

## Data Availability

Data is publicly available from the U.S. National Library of Medicine (https://clinicaltrials.gov/ct2/home)

## Declarations

### Ethical approval and consent to participate

Not applicable

### Consent for Publication

Not applicable

### Availability of data and materials

The datasets generated and/or analysed during the current study are available on the U.S. National Library of Medicine (https://clinicaltrials.gov/ct2/home).

### Authors’ contributions

FR and TP conceptualised the study, devised the methodology, and coordinated the research team. SK recruited the research team. All authors were involved in data collection and analysis, and contributed to the final manuscript.

## Acknowledgements

We are very grateful for the help of 18 research assistants that helped with compiling the data for this study: Helen Woodward, Joshua Card-Gowers, Joshua Lucas, Sultan Hussein, Mina Aries, Spatikha Sitaram, Holly Melvin, Lauren Hargreaves, Stefano Santori, Frances Kenworthy, Khalifa Saif Elyazal Ali, Maymunah Malik, Shiron Rajendran, Oliver Wright, Catherine Dominic, Holly Beckett, Tricia Tay and Daphne Lenz.

We are also thankful for the comments of Till Bruckner, Peter Grabitz, and Nicholas DeVito on the draft protocol.

## APPENDIX

**Appendix 1:**
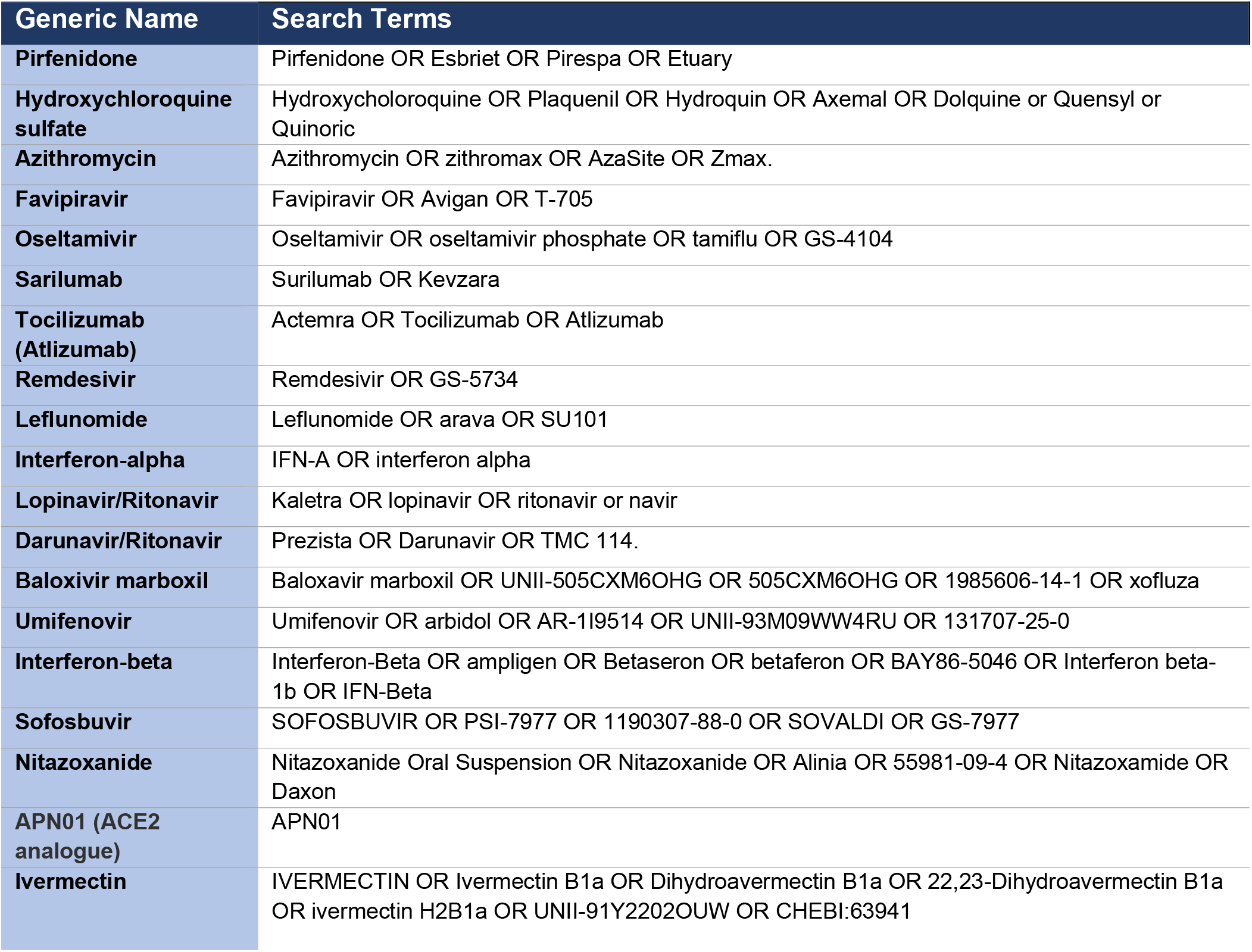
Intervention clinicaltrials.gov search terms by generic name of drug

## References

1. COVID-19 Map - Johns Hopkins Coronavirus Resource Center.

2. Xuan Y. Initial clinical results announced for favipiravir treatment of novel coronavirus pneumonia - viral clearance in four days. (Chinese). Biodiscover. 2020.

3. Wang M, Cao R, Zhang L, Yang X, Liu J, Xu M, et al. Remdesivir and chloroquine effectively inhibit the recently emerged novel coronavirus (2019-nCoV) in vitro. Vol. 30, Cell Research. Springer Nature; 2020. p. 269-71.

4. Kalil AC. Treating COVID-19 - Off-Label Drug Use, Compassionate Use, and Randomized Clinical Trials during Pandemics. JAMA - Journal of the American Medical Association. American Medical Association; 2020.

5. Sanders JM, Monogue ML, Jodlowski TZ, Cutrell JB. Pharmacologic Treatments for Coronavirus Disease 2019 (COVID-19): A Review. JAMA. 2020 Apr;

6. Vincent, Andrea L. EUA Hydroxychloroquine sulfate Health Care Provider Fact Sheet - Emergency Use Authorisation (EUA). 2020 Apr.

7. Magagnoli J, Narendran S, Pereira F, Cummings T, Hardin JW, Sutton SS, et al. Outcomes of hydroxychloroquine usage in United States veterans hospitalized with Covid-19. medRxiv. 2020 Apr;2020.04.16.20065920.

8. Borba MGS, Val FFA, Sampaio VS, Alexandre MAA, Melo GC, Brito M, et al. Effect of High vs Low Doses of Chloroquine Diphosphate as Adjunctive Therapy for Patients Hospitalized With Severe Acute Respiratory Syndrome Coronavirus 2 (SARS-CoV-2) Infection. JAMA Netw Open. 2020 Apr;3(4.23):e208857.

9. ICMJE | About ICMJE | Clinical Trials Registration.

10. FDAAA 801 and the Final Rule - ClinicalTrials.gov.

11. Bruckner T. Achieving excellence in clinical trial reporting - BIH QUEST Center report. 2020.

12. Tamiflu and Relenza: getting the full evidence picture | Cochrane.

13. Johnson RM, Doshi P, Healy D. Covid-19: Should doctors recommend treatments and vaccines when full data are not publicly available? BMJ [Internet]. 2020 Aug 24 [cited 2020 Sep 1];370:m3260. Available from: http://www.ncbi.nlm.nih.gov/pubmed/32839164

14. To Help Develop The Safest, Most Effective Coronavirus Tests, Treatments, And Vaccines, Ensure Public Access To Clinical Research Data | Health Affairs.

15. ClinicalTrialsgov. ClinicalTrials.gov Protocol Registration and Document Upload Quality Control Review Criteria. 2018.

16. Tang E, Ravaud P, Riveros C, Perrodeau E, Dechartres A. Comparison of serious adverse events posted at ClinicalTrials.gov and published in corresponding journal articles. BMC Med [Internet]. 2015 Aug 14 [cited 2020 May 28];13(1):189. Available from: http://bmcmedicine.biomedcentral.com/articles/10.1186/sl2916-015-0430-4

17. Riveros C, Dechartres A, Perrodeau E, Haneef R, Boutron I, Ravaud P. Timing and Completeness of Trial Results Posted at ClinicalTrials.gov and Published in Journals. Dickersin K, editor. PLoS Med [Internet]. 2013 Dec 3 [cited 2020 May 28];10(12):e1001566. Available from: https://dx.plos.org/10.1371/journal.pmed.1001566

18. Hartung DM, Zarin DA, Guise JM, McDonagh M, Paynter R, Helfand M. Reporting discrepancies between the ClinicalTrials.gov results database and peer-reviewed publications. Ann Intern Med. 2014 Apr 1;160(7):477-83.

19. Golder S, Loke YK, Bland M. Unpublished data can be of value in systematic reviews of adverse effects: Methodological overview [Internet]. Vol. 63, Journal of Clinical Epidemiology. Elsevier USA; 2010 [cited 2020 Sep 1]. p. 1071-81. Available from: https://pubmed.ncbi.nlm.nih.gov/20457510/

20. Golder S, Loke YK, Wright K, Norman G. Reporting of Adverse Events in Published and Unpublished Studies of Health Care Interventions: A Systematic Review [Internet]. Vol. 13, PLoS Medicine. Public Library of Science; 2016 [cited 2020 Sep 1]. Available from: https://pubmed.ncbi.nlm.nih.gov/27649528/

21. Pilkington V, Pepperrell T, Hill A. A review of the safety of favipiravir - a potential treatment in the COVID-19 pandemic? J Virus Erad. 2020;6(2):45.

22. Li G, De Clercq E. Therapeutic options for the 2019 novel coronavirus (2019-nCoV). Vol. 19, Nature reviews. Drug discovery. NLM (Medline); 2020. p. 149-50.

23. Rismanbaf A. Potential Treatments for COVID-19; a Narrative Literature Review. Arch Acad Emerg Med. 2020;8(1):e29.

24. PubChem.

25. Home - ClinicalTrials.gov.

26. WHO Statement on Public Disclosure of Clinical Trial Results Background [Internet], [cited 2020 May 28]. Available from: www.consort-statement.org

27. McHugh ML. Interrater reliability: The kappa statistic. Biochem Medica. 2012;22(3):276–82.

28. Yamey G, Gonsalves G. Donald Trump: A political determinant of covid-19. Vol. 369, The BMJ. BMJ Publishing Group; 2020.

29. Japanese flu drug “clearly effective” in treating coronavirus, says China | World news | The Guardian.

30. Positive results from initial lopinavir-ritonavir COVID-19 clinical trial.

31. PIONEER study tests treatments for mild to moderate COVID-19 — Chelsea and Westminster Hospital NHS Foundation Trust.

32. Cao B, Wang Y, Wen D, Liu W, Wang J, Fan G, et al. A Trial of Lopinavir-Ritonavir in Adults Hospitalized with Severe Covid-19. N Engl J Med. 2020 May;382(19):1787-99.

33. WMA Declaration of Helsinki - Ethical Principles for Medical Research Involving Human Subjects - WMA - The World Medical Association.

34. Martm-Martm A, Orduna-Malea E, Thelwall M, Delgado Lopez-Cozar E. Google Scholar, Web of Science, and Scopus: A systematic comparison of citations in 252 subject categories. J Informetr. 2018 Nov 1;12(4): 1160—77.

35. Schulz KF, Altman DG, Moher D. CONSORT 2010 Statement: Updated guidelines for reporting parallel group randomised trials. BMJ. 2010 Mar 27;340(7748):698–702.

36. Song F, Parekh S, Hooper L, Loke YK, Ryder J, Sutton AJ, et al. Dissemination and publication of research findings: An updated review of related biases. Health Technol Assess (Rockv). 2010;14(8):1–220.

37. Knowles RL, Ha KP, Mueller J, Rawle F, Parker R. Challenges for funders in monitoring compliance with policies on clinical trials registration and reporting: Analysis of funding and registry data in the UK. BMJ Open. 2020 Feb 17;10(2):e035283.

